# Evaluation of the Efficacy and Safety of FFX Facet Cages Compared to Pedicle Screw Fixation in Patients with Lumbar Spinal Stenosis: A Long-Term Study

**DOI:** 10.1101/2023.12.18.23300167

**Authors:** Omar Houari, Arnaud Douanla, Mehdi Ben Ammar, Mustapha Benmekhbi, Jihad Mortada, Gabriel Lungu, Cristian Magheru, Jimmy Voirin, Pablo Ariel Lebedinsky, Mariano Musacchio, Federico Bolognini, Robin Srour

## Abstract

**Objective:** The study evaluated the long-term safety and efficacy of the FFX facet cage versus pedicle screw (PS) fixation in patients with lumbar spinal stenosis (LSS).

**Methods:** A hybrid retrospective/prospective study design was used. Following a medical records review, subjects meeting the inclusion/exclusion criteria were consented and enrolled in the prospective arm of the study. CT-scans and dynamic X-rays were performed to assess fusion rates, range of motion and translation. Adverse events during the 2-year post-index procedure were also analyzed. Pre-operative and 2+ year Visual Analogue Scale (VAS) back and leg scores and Oswestry Disability Index (ODI) were also obtained.

**Results:** A total of 112 subjects were enrolled with 56 patients included in the PS and FFX groups. Mean age was 63.1±11.2 and 67.1±10.9 years and the mean number of levels operated was 1.8±0.8 and 2.3±1.0 respectively for the PS and FFX groups. There was no difference between the two groups for the primary composite fusion endpoint assessed (respectively 60.0% vs. 70.9%, p=0.120). There was also no difference in postoperative complications or adverse events during the 2-year follow-up period. A higher percentage of patients in the PS group (10.7%) required reoperation compared to the FFX group (3.6%). While both groups experienced significant improvements in VAS and ODI scores versus pre-operative assessment, there was no difference between the two groups.

**Conclusion:** The present study documents the long-term safety and efficacy of the FFX device in patients with LSS with a reduction in reoperation rate when compared to PS fixation.

## Introduction

Facet joint degeneration represents one of the most common sources of low back pain. The condition is brought upon by the loss of synovial joint space, narrowing, loss of synovial fluid and the loss of cartilage and bony overgrowth. As the facet joints degenerate over time, the resulting inflammation leads to local pain in the lumbar spine that can also radiate to the lower body.^1^

Facet joint degeneration, or facet syndrome (FS), is regarded as the most frequent form of facet pathology. Despite the usefulness and prevalence of imaging techniques for clinical back pain, it is difficult to determine an accurate rate of facet joint degeneration amongst back pain patients. For some patients, the symptoms in the setting of low back pain may lack specificity as the facet joints may mimic the pain caused by herniated discs or compressed roots. A positive diagnostic facet joint block can indicate facet joints as the chronic pain source, but the results are clouded by a high false positive rate.^2^ As such, the literature reports a wide range in the prevalence of facet joint pain.^1^ Facet joints are the primary pain generator for 10 to 15% of young adult patients with chronic low back pain with a higher rate for older population.^3^ Up to 40% of patients presenting with chronic low back pain have pain emanating from the lumbar facet joints.^4^

Facet syndrome is often present with other degenerative disorders, such as lumbar spinal stenosis (LSS). LSS refers to the narrowing in the spinal canal, in the areas of the central canal, lateral recess or the neural foramen as a result of posterior vertebral osteophyte formation, facet hypertrophy, synovial facet cysts, and/or ligamentum flavum hypertrophy. LSS is also a common source of leg and back pain. Aging, wear-and-tear changes, and traumas cause the intervertebral discs to degenerate and protrude posteriorly. As a result, the posterior elements of the vertebrae are subjected to an increased load over time. LSS presents itself as radiating pain and numbness in the lower extremity with intermittent claudication. LSS can also be secondary to degenerative spondylolisthesis, as well as rarer conditions such as space occupying lesions, post-surgical fibrosis, rheumatologic conditions, and congenital causes such as achondroplasia.^5^

The goal of LSS treatment is to reduce symptoms and improve functional status. The first line of treatment is typically conservative treatment options including physical therapy, oral anti-inflammatory medications, and epidural steroid injections.^5^ These provide short-term benefits but may fail to produce long-term improvement of pain and disability as they do not address the different origins, such as the facet joint motion or the added pressure on the nerves, that are causing the pain associated with LSS.^6^

Surgical treatment options are available for those with persisting or worsening symptoms after failed conservative treatments. Surgical treatment options aim to relieve symptoms and improve function by decompressing the neural structures that are being encroached upon. The specific treatment plan for each patient will change depending on the location of the stenosis, number of segments affected, associated deformity of spinal instability, history of previous surgery, and the surgeon’s preference. However, most LSS-related surgeries involve decompressing the spinal canal and foramina to remove the pressure factors and release the nerve roots via decompression, conventional laminectomy, or bilateral or unilateral laminotomy.^6^

While posterior fusion following decompression is intended to avoid postoperative instability, up to 10% of patients require reoperation, even those without preoperative instability.^7,8^ However, fusion itself is associated with perioperative risks, increased costs, and readmission to the hospital. In particular, lumbar pedicle screw fusion is associated with pedicle screw misplacement rates from 5 to 41% and an accelerated degeneration of the adjacent spinal segments due to the rigidity of the pedicle screw fixation systems and potential violation of adjacent facet joints during insertion.^9–11^ Additionally, up to 25% of patients undergoing laminectomy and fusion with PS experience post-fusion instability at the adjacent vertebral level to the one initially treated.^12^ PS fixation is associated with increased blood loss, soft tissue damage, and hematoma formation within the spinal canal.^13,14^ Due to these complications, post-decompression fusion is not significantly associated with lower rates of repeat surgery as compared to decompression alone.^6^

The use of facet fixation devices to achieve lumbar fusion represents an alternative to PS fixation for the treatment of patients with FS or LSS. The FFX device (SC Medica, Strasbourg, France) is an implantable facet cage designed to prevent spinal instability and facet motion by enabling lumbar facet joint fusion. This D-shaped device is made of titanium and has a serrated surface to facilitate device stabilization and is available in several sizes to ensure proper fit in the facet joint space. The implant is surgically implanted and positioned between the facet joints, with its apex oriented anteriorly (Fig. 1). Two devices are used per level with bone graft material placed inside and posterior to the implant. Previous studies have reported positive one-year outcomes associated with the placement of the FFX facet fixation device and reduced operative time and blood loss associated with the placement of the implant compared to placement of PS in patients with LSS.^15,16^ A finite element simulation comparing the device with the PS placement suggests facet fusion leads to lower mechanical loads at the adjacent levels potentially lowering the risk of adjacent segment degeneration prior to fusion while providing equivalent stability once fusion is achieved.^17^

**Figure 1.**
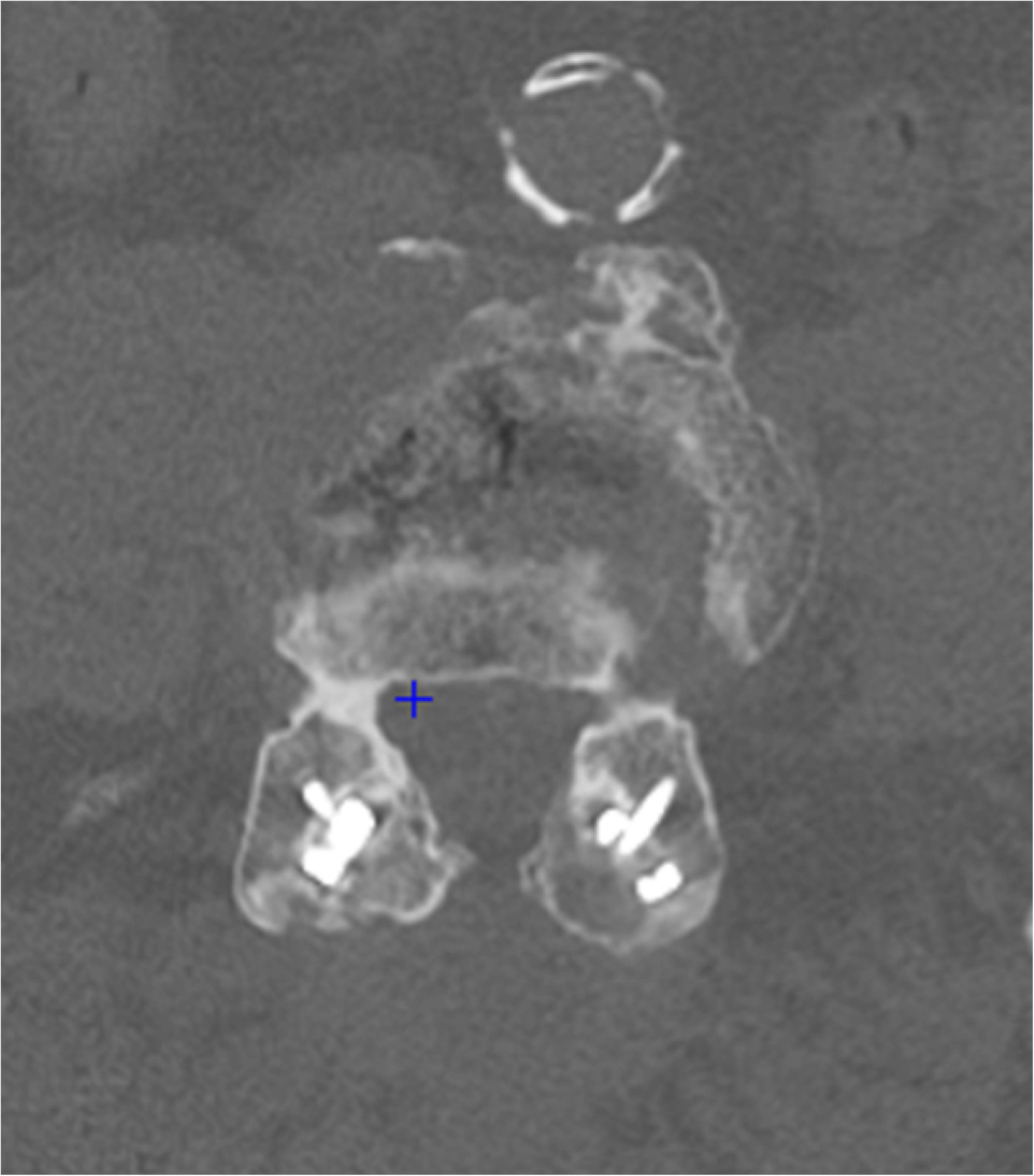
Implantation of FFX with facet screws between Facet Joints.

In order to assess the long-term performance of the FFX device and to compare these outcomes with PS fixation, we conducted a hybrid retrospective-prospective study to evaluate clinical and radiological outcomes associated with single or multi-level use of these devices.

## Material and methods

The FACETFIX study (Registration number ID-RCB 2022-A01783-40) was a single center, hybrid, retrospective and prospective study evaluating patients undergoing PS fixation or placement of FFX devices for the treatment of LSS during the defined study periods for each procedure. The study protocol was approved by an Institutional Review Board and patient consent was obtained for all subjects enrolled.

### Study Methodology

Eligible patients were identified based on a retrospective search of medical records with potential study subjects contacted and offered to participate in the study. A follow-up visit was then scheduled for patients who agreed to participate. A screening log was maintained for all eligible patients with the reasons for non-recruitment recorded.

### Inclusion/Exclusion Criteria

The retrospective component of the study corresponded to the analysis of the patients’ medical record which was conducted in two phases. The time periods for the PS and FFX groups were selected based on the use of PS as the primary implant for LSS-associated surgeries prior to the introduction of the FFX device at the study site in November 2017. The protocol stipulated that the PS group was to include all patients with LSS who underwent decompression concomitant to PS fixation (the index procedure) in reverse chronological order beginning in October 2017 back to the beginning of 2015, the date when the current IT system was implemented at the investigative site. The FFX group included all patients with LSS who underwent the index procedure with decompression concomitant to FFX fixation chronologically starting November 2017.

In addition to the above, inclusion criteria were subjects who were undergoing their first back operation for degenerative LSS greater than two years ago and at least 18 years or older at the time of the index procedure. Exclusion criteria included concomitant placement of interbody cages, unilateral PS or FFX fixation, preoperative grade ≥ II spondylolisthesis, and preoperative scoliotic deviations (Cobb angle) greater than 25°.

The prospective component of the study consisted of the follow-up of enrolled patients at a time point more than 2 years following the index procedure. Subjects received a computed tomography (CT) scan and dynamic (flexion-extension) X-rays prescription and completed a questionnaire in order to evaluate clinical outcomes. Radiographic examinations were utilized to provide objective information on the efficacy of the FFX implant to prevent spinal instability and facet motion following a decompression and spinal fusion surgery to treat lumbar spinal stenosis. Dynamic flexion-extension radiographic enables the ability to identify instabilities and to measure range of motion and translation at the treated and adjacent levels. A CT-scan was used to assess the presence of a bone bridge between the facet joints for the FFX device, or between the transverse processes or facet joints for pedicle screws.

### Study objectives and Outcomes Measurement

The primary objective of the study was to evaluate the efficacy of the FFX implant compared with pedicle screw fixation with regards to successful fusion in radiographic assessment at a time point more than 2 years following the index procedure. The criteria for successful fusion were evidence of bridging bone between the facet joints via CT-scan for both the left and right sides for all levels undergoing fixation, ≤3mm translational motion and ≤5° angular range of motion between vertebrae on flexion/extension via dynamic X-rays, and no evidence of lucency (≤25%) surrounding the device. Imaging evaluation was assessed by two specialized physicians independent from the practitioners that performed the surgery. In the event of differing evaluations, a third independent specialized physician was asked to break the tie.

Secondary objectives were to evaluate the safety of the FFX implant compared with PS fixation and to evaluate and to compare the two groups relative to improvements in Visual Analogue Scales (VAS) scores for the back and leg and disability and improvement in daily activities using the Oswestry Disability Index (ODI).

The overall safety profile was assessed based on the rate of secondary lumbar spine surgical interventions, including revisions, reoperations, or removal, or supplemental fixation during the 2-year period following the index procedure, and the number of adverse events and serious adverse events. Migration in the FFX group was defined as any displacement of the device outside the facet joint space. Misplacement was defined as any part of the device being outside the interfacet line. All subjects were evaluated for pain and disability using the VAS for back and leg pain and the ODI upon enrollment in the study. All patient assessments were conducted by independent evaluators who did not perform any of the surgeries.

### Operative Technique

PS and FFX fixation were both performed following a decompression via an open surgical approach. PS were placed using a standard placement technique with each individual investigator utilizing the PS system of their choice (CD Horizon Legacy Spine System, Medtronic; Romeo Pedicle Screw, Spineart). All pedicle screws used during the surgery were CE marked and implanted within their indications for use. For FFX placement, two implants were implanted per level. Tracking of the articular line spacing for each facet was performed with a facet chisel followed by a reviving of the facet joints with a rasp to promote fusion. Following sizing to ensure proper device fit into the joint space, the appropriately sized implant was placed on the facet holder and bone graft material was inserted into the empty space of the device. While attached to the facet holders and at the entry of the articular lines, the devices were inserted into the facet joint simultaneously on the right and left sides. The devices were then pushed into place using a supplied impactor and positioned appropriately.

### Determination of Sample Size

The primary objective of this study was to demonstrate the non-inferiority of the FFX implant versus pedicle screws regarding the primary endpoint, specifically, successful fusion at more than two years following the index procedure. A successful bony fusion rate in the FFX group of more than 86.3% was assumed based on the literature at 12 months, and 67.3 % in the PS group at 2 years.^11,13^ In the absence of a longer published follow-up for the FFX implant fusion rate, these rates were used as assumptions for more than 2 years follow-up, with the conservative assumption regarding the FFX fusion rate beyond 2 years as fusion rates increase with time post-surgery. To perform this non-inferiority analysis with a non-inferiority margin defined at 0.05, a one-sided alpha level set to 2.5 % and a 80% power; a total of 98 subjects (49 subjects by each group) will be needed to demonstrate the non-inferiority of the FFX vs. PS fixation. To reach this sample size in the per protocol population, a sample size of 116 subjects (58 subjects for each group) was needed to be enrolled in this study, to account for up to 15% of subjects to be excluded from this population due to withdrawals or major protocol deviations.

### Statistical Analysis

Quantitative variables were summarized using the following descriptive summary statistics: the number of subjects, mean, standard deviation, standard error, median, quartiles and range, minimum, and maximum value. Categorical variables (binary, nominal, and ordinal) were presented by contingency tables (frequencies and precents). The number and percentages of missing data is also be mentioned. Efficacy and safety data is presented by treatment group. Only observed data which was collected from enrolled subjects was included in the analysis.

### No subgroup analysis was conducted

Analysis of the primary fusion endpoints was conducted using a confidence interval approach with one-sided 97.5% confidence intervals (CI) used to calculate any difference in successful fusion rate between two groups (FFX and PS). The FFX group was deemed not inferior to PS group if the lower bound of the CI is above a critical value of (-0.05), which is equivalent to: if the lower limit of the confidence interval is greater than (-0.05).

Since the present study was not randomized and covariates at baseline could have theoretically been unbalanced between the two groups, a propensity score was calculated using baseline covariates that could influence the choice of the group and the primary fusion endpoint. The purpose of the propensity score was to reduce the effects of confounding factors in order to obtain an estimate of the least possible biased group effect. Group allocation and effect were considered conditionally independent of the covariates introduced in the propensity score. The variables included in the analysis included smoking status, age, sex, BMI, diabetes status, the surgeon who performed the procedure, number of levels, pre-operative pain and ODI scores and the pre-operative presence of grade 1 spondylolisthesis on at least one operated level.

Variables whose distribution is different between groups at the p<0.20 threshold were included in a multivariate model (selection=stepwise). If no variable were associated with the group allocation at the p<0.05 threshold, the propensity score was aborted.

Analyses of safety endpoints were conducted in the safety set corresponding to all patients who received FFX or PS device. Missing data was not replaced. Adverse events were summarized by relationship to the procedure, to the study device, and by seriousness. No statistical tests were performed on other variables. The VAS Leg and Back and ODI scores at more than 2 years were reported as a mean value and standard deviation for each study group.

## Results

A total of 112 patients were included in the study population. This included 56 patients in the FFX group and 56 patients in the PS fixation group. Supplemental Table 1 reports on the disposition for all patients screened for the study. As a result of an insufficient number of patients in the PS group who met the study inclusion criteria prior to the November 2017 cutoff date, additional patients who underwent PS procedures after this date were added chronologically to reach the target number of patients. This resulted in 23 of the 56 (41.1%) patients enrolled in the PS group being outside (i.e., between January 2018 and September 2020) of the original time period specified for in the study protocol. Enrollment was stopped at 112 of the 116 planned subjects to be enrolled due to a sufficient number of subjects with data achieved to meet the pre-determined number of 98 patients to demonstrate the non-inferiority of the FFX device vs. PS fixation.

Patient characteristics of the 112 patients enrolled in the study are shown in Table 1. Table 2 details procedural data related to number of levels fused, levels operated on, and number of devices implanted. A total of 111 of the 126 (88.1%) FFX devices implanted had concurrent placement of facet screws. There were no major imbalances between the two groups. Three surgeons placed FFX implants for the subjects enrolled in the study and 6 surgeons placed PS. The above included two surgeons who placed both FFX devices and PS screws for patients enrolled in the study.

**Table 1.** Patient characteristics.

| Characteristic | FFX Group<br>(n=56) | PS Group<br>(n=56) |
| --- | --- | --- |
| Sex, number (%) |  |  |
| Male | 24 (42.9) | 26 (46.4) |
| Female | 32 (57.1) | 30 (53.6) |
| Age (years); mean ± SD (range) | 67.1 ± 10.9<br>(35 – 89) | 63.1 ± 11.2<br>(31 – 84) |
| Height (cm); mean ± SD (range) | 167.9 ± 9.8<br>(148 – 190) | 165.9 ± 8.1<br>(147 – 184) |
| Weight (kg); mean ± SD (range) | 82.6 ± 18.3<br>(53 – 130) | 82.8 ± 18.4<br>(48 – 150) |
| BMI; mean ± SD (range) | 29.1 ± 4.9<br>(20 – 41) | 30.0 ± 6.0<br>(19 – 50) |
| Menopause (females only); number (%) | 25 (83.3) | 25 (83.0) |
| Osteoporosis; number (%) | 7 (12.5) | 4 (7.1) |
| Rheumatoid arthritis; number (%) | 1 (1.8) | 3 (5.4) |
| Long-term use of corticosteroids; number (%) | 3 (5.4) | 1 (1.8) |
| Insulin-dependent diabetes; number (%) | 8 (14.3) | 10 (17.9) |
| Severe arterial insufficiency or other significant peripheral vascular disease; number (%) | 8 (14.3) | 11 (19.6) |
| Permanent and complete motor, sensory or reflex deficit; number (%) | 3 (5.4) | 1 (1.8) |
| Active smoker; number (%) | 5 (8.9) | 11 (19.6) |
| Former smoker; number (%) | 12 (21.4) | 15 (26.8) |

**Table 2.** Procedural data.

| Characteristic | FFX Group<br>(n=56) | PS Group<br>(n=56) |
| --- | --- | --- |
| Number of operated levels, mean ±SD (range) | 2.3 ± 1.0<br>(1 – 5) | 1.8 ± 0.8<br>(2 – 6) |
| Level surgery performed on |  |  |
| L1-L2 | 3 | 0 |
| L2-L3 | 13 | 6 |
| L3-L4 | 37 | 25 |
| L4-L5 | 51 | 44 |
| L5-S1 | 22 | 14 |
| Number of FFX implants used, mean ±SD (range) | 2.3 ± 2.0<br>(2 – 10) | - |
| Facet screws placed, n/ FFX device implanted (%) | 111/126<br>(88.1%) | - |
| Number of pedicle screws placed, mean ±SD (range) | - | 5.6 ± 1.6<br>(4 – 10) |
| Missing data; number (%) | 1 (1.2) | 6 (10.7) |

The mean time between the initial (index) procedure, defined as the placement of the FFX implant or PS, and the 2+ year computed tomography (CT) scan, dynamic (flexion-extension) X-ray, and completion of the patient questionnaire was 3.1 + 0.9 years for the FFX group and 5.9 + 1.7 years for the PS group.

### Fusion rates

There was no statistical difference in fusion rates between the FFX and PS groups (70.9% vs. 60.0%; Chi-square test; p=0.120). The FFX group fusion rate was 75.6% for the subset of 45 patients who had concomitant facet screws implanted. The fusion rate for the FFX group was numerically greater than that of the PS group and is, therefore, not inferior to the use of pedicle screw systems (Table 3). If the more common definition of fusion was used, specifically the presence of bony bridges between the facet joints alone, the fusion rate increased to 89% and 85% for the FFX and PS groups respectively. This increased to a 91% fusion rate for the FFX group in patients who had concurrent facet screws placed with the FFX devices.

**Table 3.**
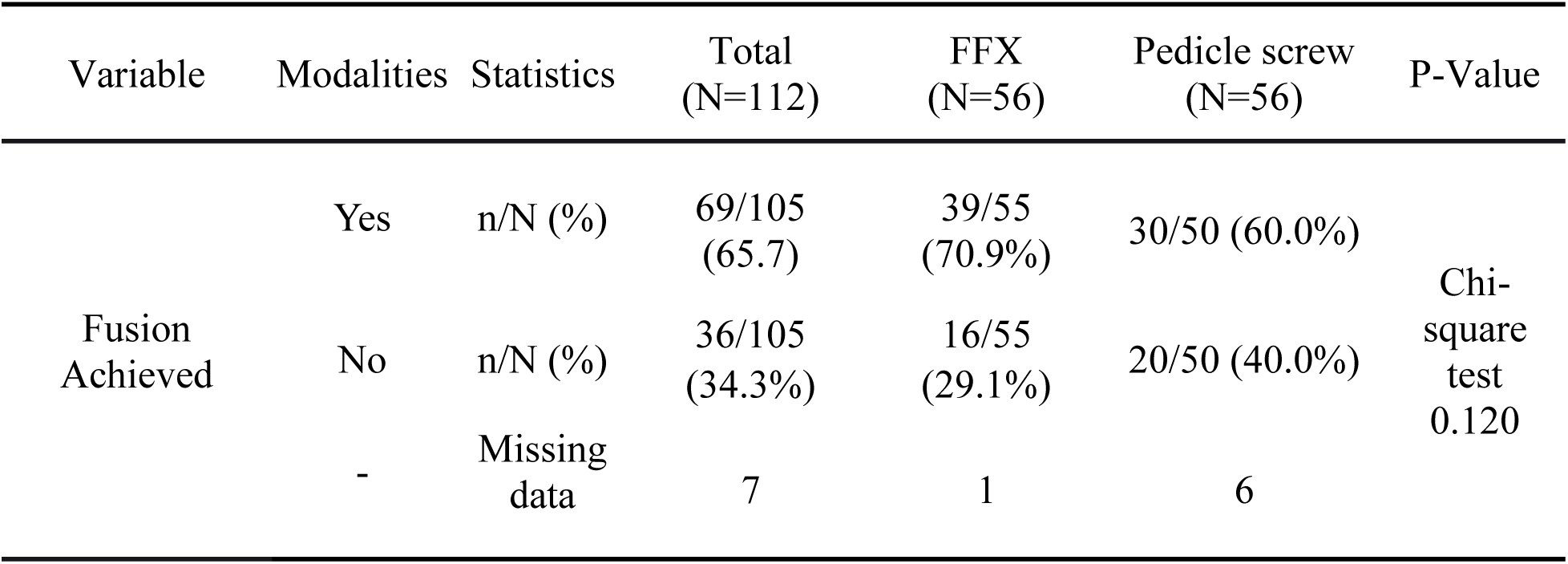
Comparison of fusion rates.

| Variable | Modalities | Statistics | Total<br>(N=112) | FFX<br>(N=56) | Pedicle screw<br>(N=56) | P-Value |
| --- | --- | --- | --- | --- | --- | --- |
| Fusion<br>Achieved | Yes | n/N (%) | 69/105<br>(65.7) | 39/55<br>(70.9%) | 30/50 (60.0%) | Chi-square<br>test<br>0.120 |
|  | No | n/N (%) | 36/105<br>(34.3%) | 16/55<br>(29.1%) | 20/50 (40.0%) |  |
|  | - | Missing<br>data | 7 | 1 | 6 |  |

A covariates analysis indicated there was no independent impact of smoking, age, sex, BMI, IDDM status, preoperative grade I spondylolisthesis, number of levels operated on, or the surgeon who performed the surgery on fusion rate. A numerical tipping point analysis to address missing data assigning missing fusion data was also conducted. Assigning the single FFX patient with missing radiographic data to the “non-fused” group resulted in a fusion rate of 69.6% and assigning the 6 patients in the PS with missing radiographic data to the “fused” group increased the fusion rate to 64.3%. These differences in fusion rates compared to the results reported in Table 3 did not change the conclusions from the original analysis. The results of the propensity score analysis also confirmed that the two cohorts were highly comparable with no differences that biased the fusion results when the FFX devices were placed with or without concomitant facet screws (Table 4).

**Table 4.**
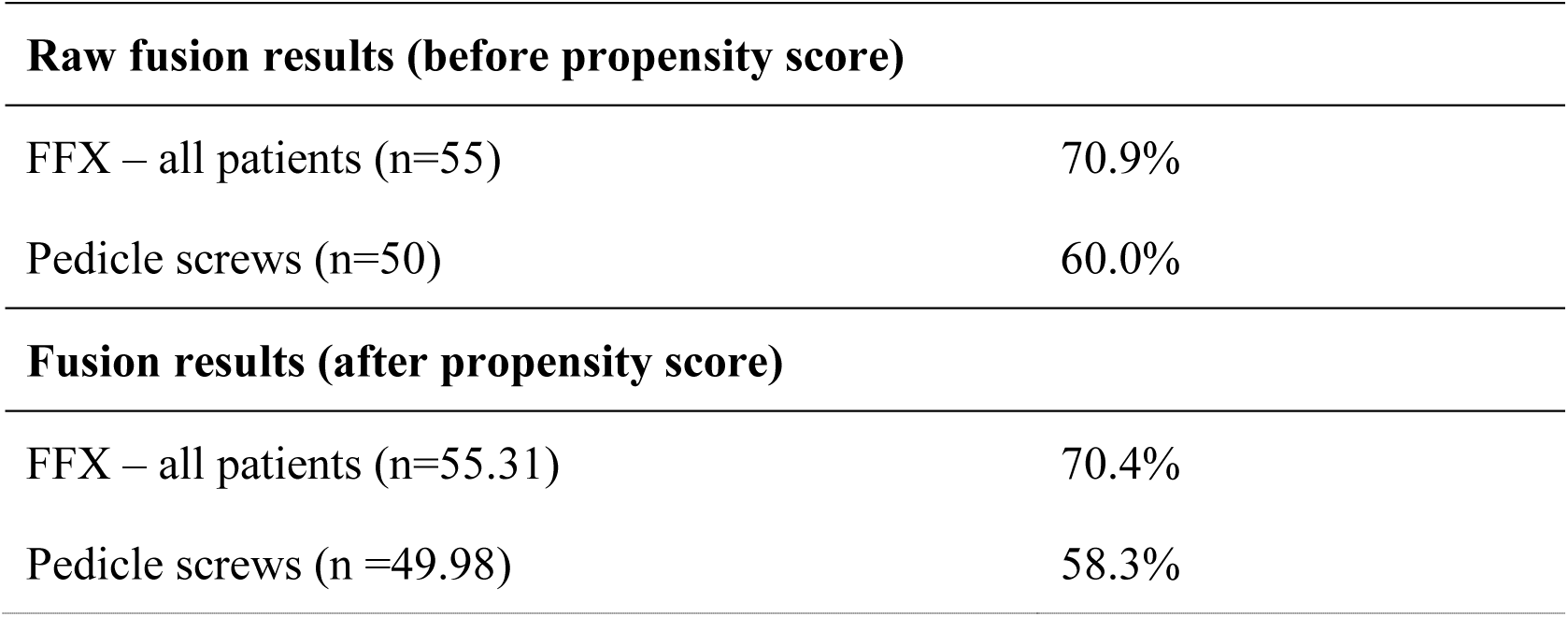
Fusion rates prior to and after propensity scoring analysis, all FFX patients.

| **Raw fusion results (before propensity score)** |  |
| --- | --- |
| FFX – all patients (n=55) | 70.9% |
| Pedicle screws (n=50) | 60.0% |
| **Fusion results (after propensity score)** |  |
| FFX – all patients (n=55.31) | 70.4% |
| Pedicle screws (n =49.98) | 58.3% |

Translation and ROM were assessed for the operative, overlying and underlying levels for both groups. The FFX group had a significantly lower translation for the underlying level compared to the PS group (0.51 ± 0.83 mm vs. 1.24 ± 1.25 mm; Wilcox test; p = 0.002). The FFX group also had a significantly lower mean ROM for the overlying level (1.59 ± 1.51 vs. 2.62 ± 2.84; Wilcox test; p = 0.024) and the underlying level (0.67 ± 1.96 vs. 1.45 ± 1.86; Wilcox test; p = 0.003) vs. the PS group. There were no significant differences in translation or ROM between the two groups for the operative levels.

Foraminal height was also measured in the FFX group and significantly increased by 3.68 ± 2.80 mm (p <0.001) at 2+ years post-surgery vs. before the surgery.

### Safety Analysis

The number of procedure-associated postoperative complications was similar between the two groups with 8 patients reporting 9 adverse events in the FFX group and 10 patients reporting 11 adverse events in the PS group (Table 5). Sciatica was the most commonly reported complication accounting for 44% of the postoperative complications in the FFX group and 73% in the PS group. No serious device-related adverse events or deaths were reported.

**Table 5.** Perioperative and Postoperative Safety Data.

| Parameter | FFX Group<br>(n=56) | PS Group<br>(n=56) |
| --- | --- | --- |
| Postoperative complications; number (%) | 9 (16.1) | 11 (19.6) |
| Sciatica | 4 (7.1) | 8 (14.3) |
| Inflammation | 2 (3.6) | 1 (1.8) |
| Cerebrospinal fluid fistula | 0 (0.0) | 1 (1.8) |
| Infection at the operative site | 1 (1.8) | 1 (1.8) |
| Hematoma not requiring surgery | 1 (1.8) | 0 (0.0) |
| Skin issue | 1 (1.8) | 0 (0.0) |
| Lumbar spine reoperation within 2 years of index procedure; number (%) | 2 (3.6) | 6 (10.7) |
| Patients reporting non-operative adverse events at 2+ year post-operative follow-up; number (%) | 39 (60.6) | 32 (57.1) |
| Number of non-operative adverse events (total) | 75 | 77 |
| Tingling | 20 | 19 |
| Numbness | 18 | 22 |
| Radiculopathy | 18 | 12 |
| Weakness | 11 | 9 |
| Limping | 5 | 12 |
| Asymmetrical weakness of motor response | 2 | 0 |
| Asymmetric decrease in reflexes | 1 | 3 |
| Device migrations; number (% of all devices placed) | 4 (1.6) | 0 (0.0) |
| Device misplacements; number (% of all devices placed) | 2 (0.8) | 3 (1.1) |

A greater percentage of patients in the PS group underwent lumbar spine reoperations during the 2-year follow-up period compared to the FFX group however, this difference was not statistically significant (10.7% vs. 3.6%; Fisher’s exact test; p=0.970). This included 2 reoperations for the FFX group and 6 for the PS group. Both of the reoperations for the FFX group were performed at L3-L4 as a result of adjacent disc degeneration. One reoperation included the placement of pedicle screws and the other did not include the placement of spinal hardware. The mean time between the index procedure and reoperation was 16.5 months (range 15 to 18 months). For the 6 patients in the PS group who had lumbar spine reoperations, three were performed at L3-L4 and one each were performed at L4-L5, L5-S1 and at both L4-L5 plus L5-S1. Four of the reoperations were due to adjacent level degeneration, one was the result of the implant being malpositioned, and one was due to a second opinion from another surgeon. Four reoperations included the placement of pedicle screws, one included the placement of FFX implants, and one did not include the placement of spinal hardware. The mean time between the index procedure and reoperation was 17.2 months (range 3 to 24 months).

Three patients in the FFX group experienced four device migration, representing 1.6% of the 252 total FFX devices placed. All 4 migrations occurred in patients who did not have facet screws concomitantly placed with the FFX device. No complications or clinical symptoms were associated with the migrations and no remedial action was required to correct. Two patients in the FFX group had two malpositioned devices, representing 0.8% of the 252 total FFX devices placed. No complications or clinical symptoms were associated with these implants and no remedial action was required to correct. Two patients in the PS group had 3 devices malpositioned, representing 1.1% of the 278 total pedicle screws placed. One patient in the PS group required reoperation resulting from a malpositioned device, but with no additional hardware placed.

### Results from Patient Feedback

Supplemental Tables 2 to 4 report the VAS Back, VAS Leg and ODI scores for the FFX and PS Group pre-operatively and at the 2+ year follow-up. While both groups experienced statistically significant improvements in all three scores versus baseline at the 2+ year follow-up, there were no statistically significant differences between the two groups at either time point for any of these functional outcome measurements (Table 6). A greater percentage of patients in the FFX group compared to the PS group were very or somewhat satisfied with their surgical management (80.4% vs. 64.3%; p=0.057).

**Table 6.** Comparison of VAS Back, VAS Leg and ODI scores at post 2-year follow-up.

| Comparison | Variable | Difference | P-value |
| --- | --- | --- | --- |
| VAS Back<br>2+ Year<br>Follow-up vs<br>Pre-op* | FFX | -2.857 | <0.001 |
|  | PS group | -2.964 | <0.001 |
| VAS Back<br>FFX vs PS<br>group† | Pre-op | -0.196 | 0.662 |
|  | 2+ Year Follow-up | -0.089 | 0.819 |
| VAS Leg<br>2+ Year<br>Follow-up vs<br>Pre-op* | FFX | -3.357 | <0.001 |
|  | PS group | -2.679 | <0.001 |
| VAS Leg<br>FFX vs PS<br>group | Pre-op | 0.286 | 0.587 |
|  | 2+ Year Follow-up | -0.393 | 0.376 |
| ODI<br>2+ Year<br>Follow-up vs<br>Pre-op* | FFX | -0.170 | <0.001 |
|  | PS group | -0.064 | 0.013 |
| ODI<br>FFX vs PS<br>group | Pre-op | 0.065 | 0.035 |
|  | 2+ Year Follow-up | -0.041 | 0.260 |

\*Dunnett adjustment †Tukey adjustment
VAS = Visual Analogue Scale; ODI = Oswestry Disability Index

## Discussion

Pedicle screw fixation following decompression is currently considered the standard technique for achieving fusion and spinal stability in patients with lumbar spinal stenosis.^18^ The present study demonstrated that the FFX implant can achieve similar or greater fusion rates than with pedicle screws without increased procedural risks. These results provide further long-term evidence documenting the safety and efficacy of the device, building on previously reported 1 year device outcomes.^15^ The data reported from this study also demonstrates an improvement in functional outcomes in patients receiving FFX implants as measured by VAS and ODI, with a high level of patient satisfaction with their surgical outcomes.

The FFX device provides several potential advantages to the use of pedicle screw fixation systems in patients with LSS receiving decompression where postsurgical spinal stabilization is desired. The less rigid fixation and reduced load projects with the FFX device compared to pedicle screws can result in less adjacent segment degeneration and theoretically a reduced need for subsequent surgical procedures.^17^ Additionally, the placement of the FFX device does not inhibit the ability to perform a fusion procedure with pedicle screw placement in the future, if desired.

While there was a low misplacement rate with the FFX device in the present study, this compared favorably to the reported misplacement rate for pedicle screws. Additionally, even though there were instances of device migration in this study, FFX device migration generally results in the device ending up in the muscle, a significant advantage versus the risks associated with pedicle screw misplacement in the canal and the potential need for reoperation to avoid permanent injuries or disabilities. Of note is that all 4 device migrations reported in the present study occurred in patients who did not have a concomitant facet screw placed with the device.

Placement of pedicle screws via open lumbar surgery using a posterior approach is associated with significant soft tissue damage and blood loss.^19,20^ Procedural associated blood loss with PS placement increases the risk of post-operative infections, hematoma formation within the spinal canal, and blood transfusion.^21,22^ The use of PS constructs can also result in adjacent level degeneration due to the rigidity produced by this approach and the resultant overload of anatomical structures.^23^ Since the FFX device can be placed under direct visualization, the procedure avoids the use of fluoroscopy and associated radiation risks that are required for pedicle screw placement. Additionally, the reduced surgical exposure required for the procedure and simpler placement technique compared to the pedicle screw fixation systems translates to the potential for a reduction in operative time and blood loss for patients. A recent non-randomized, retrospective study compared mean operative time and estimated blood loss between patients undergoing posterior lumbar fusion surgery for LSS with either pedicle screw fixation or the FFX device.^16^ The author reported that PS fixation was associated with significantly longer mean operative time compared to placement of the FFX device (152.5 ± 39.4 vs. 99.4 ± 44.0 minutes; p<0.001). Mean operative blood loss was also significantly greater for PS vs. FFX procedures (446.5 ± 272.0 vs. 251.0 ± 315.9 mL; p<0.001). These differences were independent of the number of levels operated on.

There are several potential limitations associated with the present study. This includes the non-randomized study design, and the enrollment of patients from a single center even though the study included patients from a total of 7 surgeons. Additionally, the nearly 3-year difference between the two groups relative to when CT-scans were performed was potentially favorable to the PS fixation group in terms of fusion rate. Exclusion of patients in whom lumbar cages were placed also limits the ability to translate these results to patients where cages are used in combination with PS fixation or implantation of the FFX device.

## Conclusion

The results of the present study document the safety and efficacy of the FFX device over a 2-year follow-up period in patients with lumbar spinal stenosis and demonstrate a favorable comparison to PS placement for several outcome parameters. Clinical outcomes included a reduction in both pain and disability following surgery with a high fusion rate and a low reoperation rate. Additional studies are needed to assess the long-term results of facet fixation and to directly compare this approach with other procedures and fixation constructs.

## Data Availability

All data produced in the present study are available upon reasonable request to the authors.

## Acknowledgements

Statistical analysis support for the present study was provided by Soladis Clinical Studies, Roubaix, France. Medical writing assistance was provided by Larry Yost (The Atticus Group, Portsmouth, New Hampshire, USA).

