## Supplemental Tables for "Evaluation of the Efficacy and Safety of FFX Facet Cages Compared to Pedicle Screw Fixation in Patients with Lumbar Spinal Stenosis: A Long-Term Study"

**Supplemental Materials**

**Supplemental Table 1.** Screening Log

| | FFX Group | PS Group |
|---|---|---|
| Number of patients identified via medical records search | 131 | 101 |
| Number of patients screened | 84 | 75 |
| Number of screened patients who were not enrolled in study | 28 | 19 |
| Reason for non-enrollment | | |
| - Did not meet all eligibility criteria | 11 | 4 |
| - Count not be reached | 4 | 3 |
| - Refused to participate | 6 | 6 |
| - Other | 7 | 6 |
| Total patients enrolled in study | 56 | 56 |

**Supplemental Table 2.** VAS Back Scores Descriptive Data – Pre-Op and Post 2 Year Follow-up

| Variable | Statistics | Total (N=112) | FFX (N=56) | PS group (N=56) |
|---|---|---|---|---|
| Pre-op | Number of data (missing data) | 112 (0) | 56 (0) | 56 (0) |
| | Mean (± Std) | 6.40 (±2.36) | 6.30 (±2.69) | 6.50 (±2.00) |
| | Median (Q1 ; Q3) | 7.00 (6.00;8.00) | 7.00 (6.00;8.00) | 7.00 (6.00;8.00) |
| | (Min ; Max) | (0.00;10.00) | (0.00;10.00) | (0.00;9.00) |
| 2+ Year Follow-up | Number of data (missing data) | 112 (0) | 56 (0) | 56 (0) |
| | Mean (± Std) | 3.49 (±2.05) | 3.45 (±2.17) | 3.54 (±1.93) |
| | Median (Q1 ; Q3) | 3.00 (2.00;5.00) | 3.00 (2.00;5.00) | 3.00 (2.00;5.00) |
| | (Min ; Max) | (0.00;8.00) | (0.00;8.00) | (0.00;8.00) |

**Supplemental Table 3.** VAS Leg Scores Descriptive Data - Pre-Op and Post 2 Year Follow-up

| Variable | Statistics | Total (N=112) | FFX (N=56) | PS group (N=56) |
|---|---|---|---|---|
| Pre-op | Number of data (missing data) | 112 (0) | 56 (0) | 56 (0) |
| | Mean (± Std) | 5.59 (±2.77) | 5.73 (±2.82) | 5.45 (±2.73) |
| | Median (Q1 ; Q3) | 6.00 (4.50;7.50) | 6.00 (4.50;8.00) | 6.00 (4.50;7.00) |
| | (Min ; Max) | (0.00;10.00) | (0.00;10.00) | (0.00;10.00) |
| 2+ Year Follow-up | Number of data (missing data) | 112 (0) | 56 (0) | 56 (0) |
| | Mean (± Std) | 2.57 (±2.34) | 2.38 (±2.37) | 2.77 (±2.30) |
| | Median (Q1 ; Q3) | 2.00 (0.00;4.00) | 2.00 (0.00;4.00) | 3.00 (0.00;4.00) |
| | (Min ; Max) | (0.00;10.00) | (0.00;10.00) | (0.00;8.00) |

**Supplemental Table 4.** ODI Scores Descriptive Data – Pre-Op and Post 2 Year Follow-up

| **Variable** | **Statistics** | **Total (N=112)** | **FFX (N=56)** | **PS group (N=56)** |
|---|---|---|---|---|
| Pre-op | Number of data (missing data) | 111 (1) | 56 (0) | 55 (1) |
| | Mean (± Std) | 0.49 (±0.16) | 0.52 (±0.16) | 0.46 (±0.16) |
| | Median (Q1 ; Q3) | 0.51 (0.36;0.62) | 0.54 (0.40;0.64) | 0.44 (0.33;0.58) |
| | (Min ; Max) | (0.12;0.86) | (0.12;0.86) | (0.12;0.78) |
| 2+ Year Follow-up | Number of data (missing data) | 111 (1) | 56 (0) | 55 (1) |
| | Mean (± Std) | 0.37 (±0.19) | 0.35 (±0.19) | 0.39 (±0.20) |
| | Median (Q1 ; Q3) | 0.38 (0.22;0.53) | 0.37 (0.20;0.51) | 0.42 (0.26;0.56) |
| | (Min ; Max) | (0.00;0.80) | (0.00;0.64) | (0.00;0.80) |
